# From text to tables: Zero-shot extraction of structured clinical data from free-text CT scan reports using foundational large language models

**DOI:** 10.1101/2025.10.05.25337366

**Authors:** Alex Hongslo, Amulya Gupta, Quynh Nguyen, Jake Caldwell, Ben Choi, Christopher J. Harvey, Jeffrey Thompson, Diego Mazzotti, Zijun Yao, Amit Noheria

## Abstract

**Background:** Large language models (LLMs) are being explored for multiple applications in medical research, including medical text classification. We evaluate the performance of 5 off-the-shelf LLMs for classifying free-text CT angiography reports for pulmonary embolism (PE)- related diagnostic labels.

**Methods:** We assessed 1,025 manually labeled CT reports using 5 LLMs (ChatGPT-4o, Llama 3.3 70b, Llama 3.1 8b, Llama 3.2 3b, Mixtral 8×7b) with zero-shot prefix prompts. Labels included acute PE, bilateral PE, and large PE. Voting ensemble models combining multiple LLM outputs were also tested.

**Results:** Llama 3.3 70b and ChatGPT-4o outperformed smaller models for all classification tasks. Highest accuracies were 96.6% (acute PE), 92.7% (bilateral PE), and 82.6% (large PE). Voting ensemble models offered no or minimal improvement in classification performance.

**Conclusions:** Off-the-shelf LLMs, particularly larger ones, can classify free-text reports with high accuracy using simple prompts. Further work is needed to optimize prompting strategies and evaluate hybrid approaches.

## INTRODUCTION

Recently developed large language models (LLMs) are being explored for multiple potential applications in healthcare and medicine, including decision aids systems, clinical encounters summarization, patient-facing chatbots, administrative tasks, literature review, and others (1).

In clinical research, extraction of structured text from free-text medical reports is another use-case for LLMs. Retrospective studies in medicine often rely on the presence of datasets with structured labels for exposures and outcomes to facilitate quantitative analysis. Frequently, this entails significant human effort devoted to tedious manual chart reviews which can be a bottleneck for sample size and statistical power. This becomes even more pertinent in the context of machine learning for clinical prediction tasks, where the requirement for training data can run into hundreds of thousands of records.

Historically, various natural language processing (NLP) systems have been considered the standard for automated text classification and information extraction from clinical free text (2). Although these systems are relatively computationally efficient, they lack flexibility and require substantial task-specific customization and pre-processing steps (3). In contrast, LLMs offer greater semantic flexibility and contextual understanding; however, they are computationally intensive, often produce hallucinated outputs, and may not consistently adhere to prompt instructions (4).

Given these considerations, we investigated the utility of commonly available, off-the-shelf LLMs for automating the extraction of pulmonary embolism (PE) labels from free-text computed tomography angiography (CTA) reports, using human-annotated labels as the reference standard.

## METHODS

This study was conducted under the approval of the institutional review board of the University of Kansas. We identified CTA reports from the University of Kansas Health System (2010–2022) using a natural language search for PE-related terms in Illuminate® Insight™ (Softek Illuminate Inc., Overland Park, KS), as part of a larger project on PE diagnosis and outcomes. Reports were randomly assigned to 4 authors for chart review and manual classification via an extensive structured REDCap® form.

The anonymized CTA reports were uploaded to a HIPAA-compliant cloud-based institutional digital research platform. Five LLM endpoints—ChatGPT-4o, Llama 3.3 (70 billion parameters), Llama 3.1 (8 billion parameters), Llama 3.2 (3 billion parameters), and Databricks Mixtral (8×7 billion parameters)—were independently queried with similar prompts to classify CTA reports for acute PE (Yes/No) (**Table 1**). Within positive cases, labels for bilateral PE and large PE (defined as lobar involvement or greater) were also queried. While the LLMs were explicitly prompted to produce single word output, regardless, 10-30% outputs of the smaller LLMs (i.e. Llama 8b, Llama 3b, and Mixtral 8×7b) were multi-word sentences. These were post-processed using simple string-matching rules to extract binary labels. For each classification task, we constructed 2 voting ensemble models: one by aggregating the binary outputs from all 5 LLMs, and the other using only the 2 best-performing LLMs. The ensembles were evaluated at varying thresholds of number of votes to assess performance.

**Table 1.** Prompts provided to the large language model for each report.

| Task | Prompt |
| --- | --- |
| Acute PE (Yes/No) | After reading the following CT scan report of a single patient with suspected PE, answer "Yes" or "No" whether the patient's reports strongly suggest acute pulmonary embolism. Make sure to only return a single word answer, "Yes" or "No" without any explanations, words, symbols: |
| Bilateral PE (Yes/No) | After reading the following CT scan report of a single patient with acute pulmonary embolism (PE), answer "Yes" or "No" whether the patient's report strongly suggests that acute PE is bilateral i.e. acute PE findings are present in both the lungs. Make sure to only return a single word answer, "Yes" or "No" without any explanations, words, symbols: |
| Extent of PE (Large/Small) | After reading the following single CT scan report of a patient with acute pulmonary embolism (PE), determine if the embolism is large, defined as affecting the lobar arteries, right or left pulmonary arteries, or being a saddle PE. The embolism should be classified as small if it is segmental or sub-segmental. If the affected arteries are not provided but the report categorically mentions it to be small, then answer it to be small. In case findings are suggestive of both small and large PE, answer it to be large. Make sure to only answer as a single word, "Small" or "Large" without any explanations, letters, words, or symbols: |

The performance of LLMs was compared using sensitivity, specificity, precision, accuracy, F1-score, and single-point area under the receiver operating characteristic (AUC) curves with human-labeled data serving as the ground truth. Data analysis was performed in JMP Pro 17 (SAS institute, Cary, NC) and R version 4.4.1 (http://www.r-project.org/).

## RESULTS

A total of 1,106 CTA reports were manually reviewed for diagnosis of acute PE and 81 of these reports were excluded from analysis due to equivocal or undetermined findings. The remaining 1,025 reports (54% female, 59.3 ± 15.8 years age, 62% white) were utilized for analysis with 660 (64.4%) indicating the presence of an acute PE, of which 300 (46.7%) were bilateral, and 187 (28.3%) were large PE. Manually extracted Yes/No labels for bilateral PE were missing in 18 (2.7%) patients, and these were excluded from the analysis.

### Acute PE (Yes/No)

As shown in **Table 2**, Llama 3.3 70B had the highest accuracy (96.6%), followed by ChatGPT-4o (95.7%) and Mixtral 8×7b (95.7%). While the smaller models, i.e. Llama 3.1 8b and Llama 3.2 3b had lower accuracies (90.5% and 87.9% respectively), their sensitivities were relatively high (98.9% and 96.7% respectively).

**Table 2.** Performance of large language models in CTA report classification tasks for pulmonary embolism labels.

| Classification Task and Model | Sensitivity (%) | Specificity (%) | Precision (%) | Accuracy (%) | F1-score (%) |
| --- | --- | --- | --- | --- | --- |
| **Acute PE** |  |  |  |  |  |
| ChatGPT-4o | 94.6 | 97.8 | 98.7 | 95.7 | 96.6 |
| Llama 3.3 70b | 95.6 | 98.4 | 99.1 | 96.6 | 97.3 |
| Llama 3.1 8b | 98.9 | 75.3 | 87.9 | 90.5 | 93.1 |
| Llama 3.2 3b | 96.7 | 72.1 | 86.2 | 87.9 | 91.1 |
| DBRX Mixtral 8x7b | 95.9 | 95.3 | 97.4 | 95.7 | 96.6 |
| **Bilateral acute PE** |  |  |  |  |  |
| ChatGPT-4o | 98.7 | 87.1 | 87.1 | 92.5 | 92.5 |
| Llama 3.3 70b | 98.3 | 87.7 | 87.5 | 92.7 | 92.6 |
| Llama 3.1 8b | 99.3 | 20.8 | 52.4 | 57.5 | 68.6 |
| Llama 3.2 3b | 88.6 | 42.5 | 57.6 | 64.1 | 69.8 |
| DBRX Mixtral 8x7b | 95.0 | 76.0 | 77.7 | 84.9 | 85.5 |
| **Large PE** |  |  |  |  |  |
| ChatGPT-4o | 87.7 | 80.6 | 64.1 | 82.6 | 74.0 |
| Llama 3.3 70b | 85.0 | 78.4 | 60.9 | 80.3 | 71.0 |
| Llama 3.1 8b | 96.2 | 24.1 | 33.3 | 44.5 | 49.5 |
| Llama 3.2 3b | 77.0 | 44.6 | 35.5 | 53.8 | 48.6 |
| DBRX Mixtral 8x7b | 91.4 | 55.0 | 44.5 | 65.3 | 59.9 |

### Bilateral PE (Yes/No)

Accuracy for classifying bilateral PE was highest for Llama 3.3 70b (92.7%) which was closely followed by ChatGPT-4o (92.5%). However, the performance of the other 3 models was poor with Llama 3.1 8b having the lowest accuracy of just 57.5%. While all the models had good sensitivity, the specificities were relatively lower with the best performing model, i.e. Llama 3.3 70b, having a specificity of 87.1%.

### Extent of PE (Large/Small)

Accuracies were lowest for large PE diagnosis amongst the three classification tasks. ChatGPT-4o was the best performing LLM with accuracy of 82.6%, followed by Llama 3.3. 70b (80.3%) and Mixtral 8×7b (65.3%). Both sensitivity and specificity were moderate for ChatGPT-4o (87.7% and 80.6% respectively) and Llama 3.3 70b (85.0% and 78.4% respectively). The comparative performance of the models for the classification tasks can be visualized as single-point AUCs, as shown in **Figure 1**.

**Figure 1.**
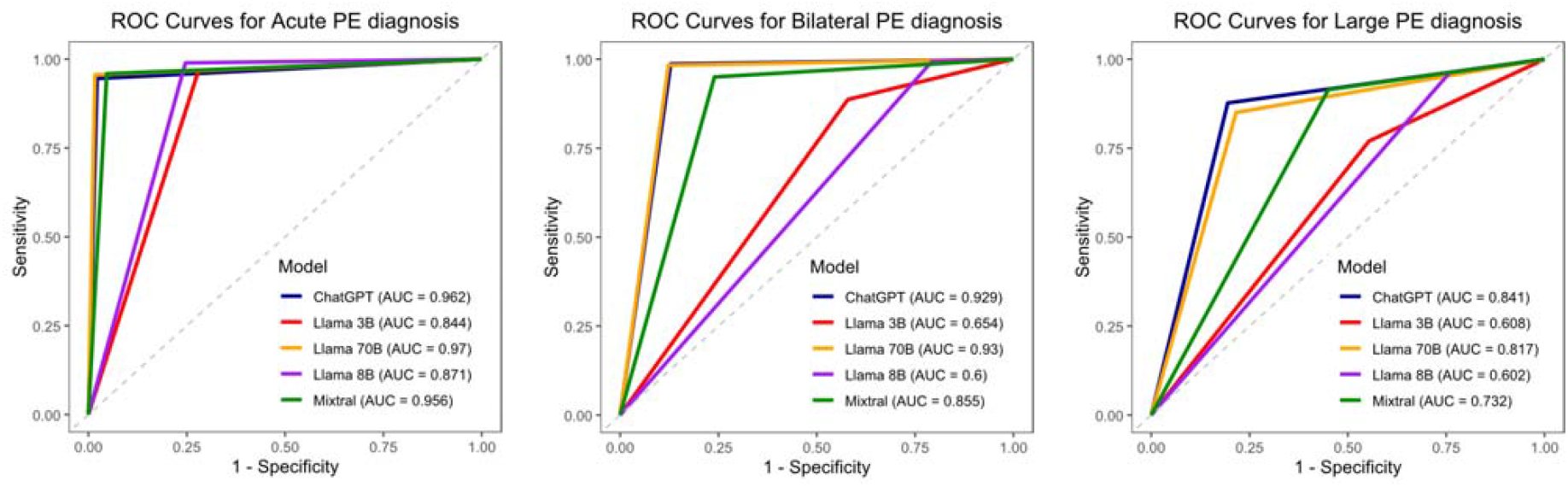
Single-point area under the receiver operating characteristic (AUC) curves of classification performance of large language models for acute PE (left), bilateral PE (center), and large PE (right).

As shown in **Supplementary Tables 1 and 2**, creation of voting ensemble models by combining outputs led to modest or no increase in accuracies. For the 5 LLM voting ensemble, the highest accuracies and vote thresholds were: 96.7% for acute PE (≥3 votes), 92.5% for bilateral PE (≥4 votes), and 82.4% for large PE (≥5 votes). Similarly, for the 2 LLM voting ensemble combining ChatGPT-4o and Llama 3.3 70b outputs, the highest accuracies and vote thresholds were: 96.7% for acute PE (≥1 votes/ OR operator), 93.5% for bilateral PE (at 2 votes/ AND operator), and 82.3% for large PE (at 2 votes/ AND operator)

## DISCUSSION

We evaluated the performance of 5 publicly available, off-the-shelf LLMs in extracting PE-related diagnostic labels from free-text CT scan reports. The key findings of our study are as follows. First, all tested models, including smaller and computationally efficient LLMs such as Llama 3.1 8b, demonstrated high accuracy for the relatively straightforward task of classifying acute PE. Second, performance declined across all models for more complex classifications, such as identifying bilateral PE or large PE. In particular, the smaller LLMs (Llama 3.2 3b and Llama 3.1 8b) performed very poorly on these tasks, indicating limited utility for more nuanced clinical extractions. Third, voting ensemble models that aggregated binary outputs from multiple LLMs did not yield substantial improvements in overall performance. However, adjusting voting thresholds within these ensembles may allow for tuning sensitivity or specificity, depending on the clinical requirements of the task.

This analysis builds upon the several recent studies evaluating LLMs for structuring free-text clinical data (5-9). As identified in these studies, although LLMs are relatively easy to deploy and often achieve high accuracies, they suffer from unpredictable inconsistencies that are hard to predict and even more difficult to systematically mitigate (10). For instance, during our analysis, some LLMs sporadically generated multi-word sentences instead of Yes/No responses. Further, LLMs are prone to hallucinations, where they generate labels when information is non-existent or ambiguous (11).

As such, LLMs are generally built on transformer architectures, which are not specifically designed for information extraction but for predicting the next word in a sequence based on contextual embeddings. These models rely on a self-attention mechanism, which allows each word in the input to be dynamically weighted in relation to every other word, enabling the model to capture complex contextual information (12). Consequently, LLMs do not explicitly parse each word to identify predefined terms associated with a particular classification. Instead, they generate responses based on the representation of the input as encoded in the embedding space, making their outputs less transparent and more probabilistic in nature. Moreover, in most LLM endpoints, the prompt (instructions) and input text (e.g., clinical reports) are ultimately concatenated into a single input sequence. This may make the models susceptible to irregularities in the free text itself, such as escape characters, formatting artifacts, or stray phrases that may inadvertently resemble instructions.

Regardless, as demonstrated in our study, LLMs show good empirical performance in zero-shot free-text classification, especially for the straightforward tasks like acute PE diagnosis. Based on our observations, we have presented a general outline for binary label classification methodology in **Figure 2**.

**Figure 2.**
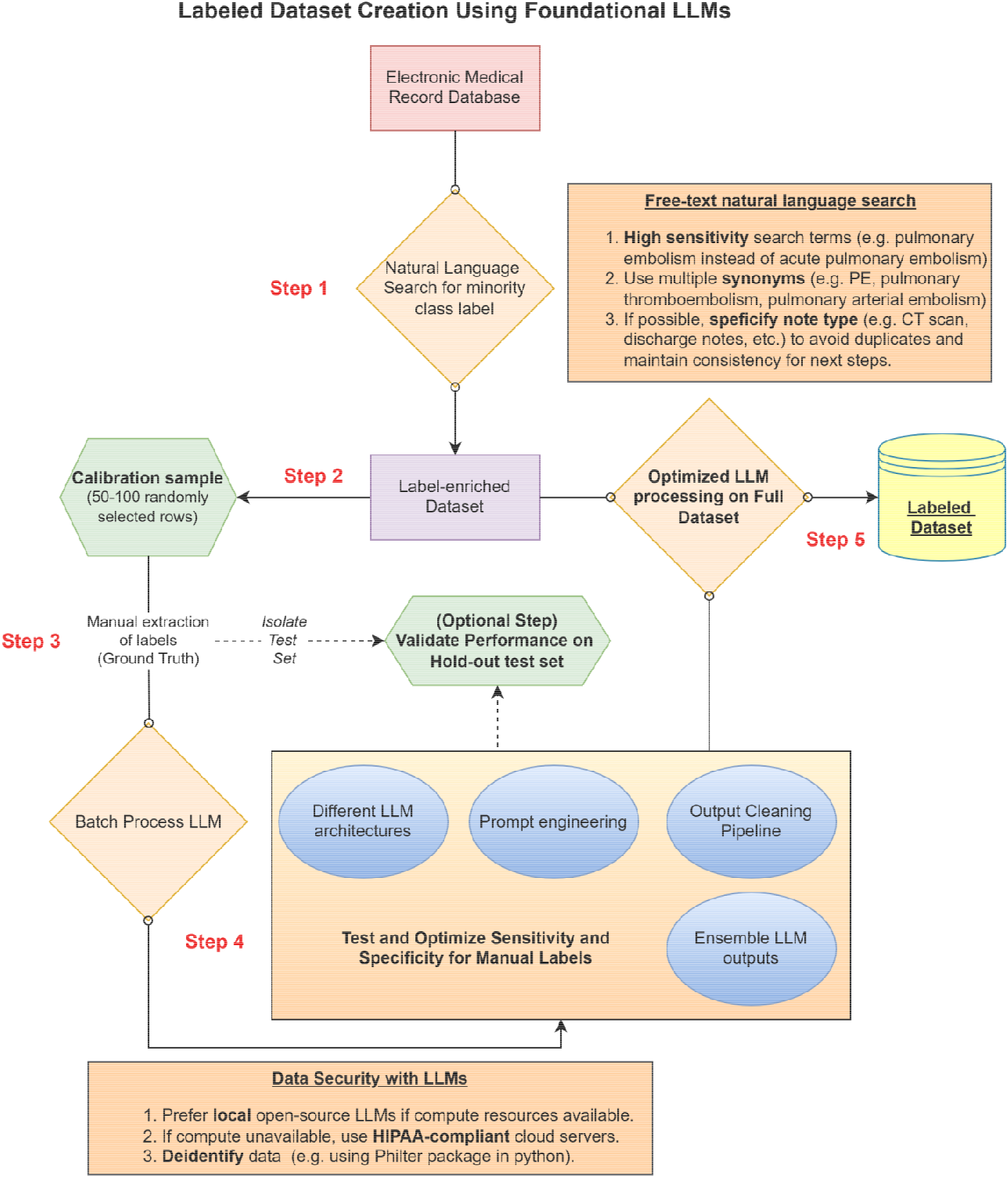
A general outline for binary label classification of free-text clinical reports using a simple large language model and natural language search framework.

Alternatives to LLMs include rule-based NLP, where manually specified explicit rules and pattern matching are used to classify free-text reports. However, these are brittle to minor variations in language, and therefore, may demonstrate low sensitivity or specificity (13,14). Machine learning NLP and hybrid NLP are other approaches to text classification, which can achieve high accuracies but require large number of training samples and extensive testing before deployment (15,16). Fine-tuning LLMs can improve classification performance but is extremely resource-intensive, requires large labeled datasets, and produces task-specific models unsuitable for smaller-scale projects (17). Another framework is Retrieval-Augmented Generation (RAG), where relevant passages from large data sources are retrieved and fed to an LLM to enable generation grounded in real-world data. RAG can be used to extract labels reflecting patient health states by leveraging the broader context of the electronic medical record, rather than relying solely on isolated reports from specific modalities (18). While we primarily used simple prefix prompts in this study, several other prompting techniques have been proposed for improving LLM yield (19). Chain-of-thought prompting is an important technique where the model is prompted to generate intermediate reasoning steps, which may improve classification accuracy and increase transparency for data point extraction (20).

### Limitations

This analysis has several limitations. First, we used label-enriched reports that were pre-filtered using natural language search for PE-related terms. As a result, LLM performance observed in this study may not generalize to unfiltered, general CT scan reports. Second, each case was manually labeled by a single reviewer from a group of four, without cross-verification. This may have introduced labeling errors and inter-observer variability. Finally, we did not systematically evaluate the impact of model parameters (e.g., temperature) or prompt variations on LLM performance, which could influence output consistency and accuracy.

## CONCLUSIONS

With appropriate validation and post-processing, off-the-shelf LLMs can effectively classify free-text CT scan reports into diagnostic labels using simple zero-shot prompts. Larger, more computationally intensive models (such as Llama 3.3 70b and ChatGPT-4o) consistently outperform smaller models (such as Llama 3.1 8b and Llama 3.2 3b), particularly in more nuanced classification tasks. Further research is warranted to explore the utility of LLMs in medical text classification across diverse prompting strategies, model architectures, and hybrid approaches that integrate traditional NLP techniques.

## Data Availability

The data underlying this study contains Protected Health Information (PHI) and cannot be shared. Access is restricted to individuals authorized by the Institutional Review Board in accordance with institutional guidelines.

## Notes

### Competing Interest Statement

The authors have declared no competing interest.

### Funding Statement

None

### Author Declarations

IRB of University of Kansas gave ethical approval for this work.

